# Clinical Associations of Functional Dyspepsia with Gastric Dysrhythmia on Electrogastrography: A Comprehensive Systematic Review and Meta-Analysis

**DOI:** 10.1101/2021.01.19.21250140

**Authors:** C Varghese, DA Carson, S Bhat, TCL Hayes, AA Gharibans, CN Andrews, G O’Grady

**Affiliations:** Department of Surgery, Faculty of Medical and Health Sciences, The University of Auckland, Auckland 1023, New Zealand; Department of Medicine, University of Calgary, Calgary, Canada; Auckland Bioengineering Institute, The University of Auckland, Auckland 1010, New Zealand

**Keywords:** Biomarkers, Gastric Electrical Activity, Electrogastrography, Slow Waves, Gastroduodenal disorders, Functional dyspepsia

## Abstract

**Background:** Functional dyspepsia (FD) is a common gastroduodenal disorder, yet its pathophysiology remains poorly understood. Bioelectrical gastric slow wave abnormalities are thought to contribute to its multifactorial pathophysiology. Electrogastrography (EGG) has been used to record gastric electrical activity, however the clinical associations require further evaluation.

**Aims:** This study aimed to systematically assess the clinical associations of EGG in FD.

**Methods:** MEDLINE, EMBASE, and CENTRAL databases were systematically searched for articles using EGG in adults with FD. Primary outcomes were percentage normal *vs* abnormal rhythm (bradygastria, normogastria, tachygastria). Secondary outcomes were dominant power, dominant frequency, percentage coupling and the meal responses.

**Results:** 1751 FD patients and 555 controls from 47 studies were included. FD patients spent less time in normogastria while fasted (SMD −0.74; 95%CI −1.22 - −0.25) and postprandially (−0.86; 95%CI −1.35 - −0.37) compared to controls. FD patients also spent more fasted time in bradygastria (0.63; 95%CI 0.33 – 0.93) and tachygastria (0.45; 95%CI 0.12 – 0.78%). The power ratio (−0.17; 95%CI −0.83 - 0.48), and dominant frequency meal-response ratio (0.06; 95%CI −0.08 - 0.21) were not significantly different to controls. Correlations between EGG metrics and the presence and timing of FD symptoms were inconsistent. EGG methodologies were diverse and variably applied.

**Conclusion:** Abnormal gastric slow wave rhythms are a consistent abnormality present in FD, as defined by EGG, and therefore likely play a role in pathophysiology. The aberrant electrophysiology identified in FD warrants further investigation, including into underlying mechanisms, associated spatial patterns, and symptom correlations.

## Introduction

Functional dyspepsia (FD) is the most common of the gastroduodenal functional disorders, being defined by the Rome IV Criteria as one or more of: postprandial fullness, early satiety, epigastric pain or burning, occurring regularly, and not explained by routine clinical investigations ^1^. FD affects up to 16% of the general population ^2^, with a near tripling in the incidence of the postprandial distress syndromes subtype over the last two decades ^3^.

Despite this substantial impact, the underlying pathophysiology of FD is poorly understood. The disorder is likely heterogenous, with putative mechanisms including: brain-gut axis dysfunction, visceral hypersensitivity and alterations in immune, gut microbiome, and mucosal function ^24–7^. Abnormal gastric electrical activity is also thought to be contributory and has historically been measured using electrogastrography (EGG). However, the clinical associations of EGG recordings are poorly defined and incompletely understood.

The recent emergence of new techniques for recording gastric electrophysiology, such as high-resolution and body surface mapping approaches, and their promising symptom correlations, motivates renewed interest in the electrophysiology of functional gastroduodenal disorders ^8–10^. Investigating this aberrant slow wave activity could yield biomarkers capable of informing personalized diagnosis and management in a subset of FD patients ^8,11^.

Many previous studies have evaluated gastric electrophysiology in FD using EGG, providing an important foundation for informing the application of new techniques. The aim of this study was therefore to systematically review and meta-analyse the existing clinical associations between EGG recordings in patients with FD, in order to synthesise known pathophysiology and guide future studies.

## Methods

This systematic review was conducted in accordance with the Meta-Analysis of Observational Studies in Epidemiology (MOOSE) guidelines for systematic reviews and meta-analysis of observational studies ^12^.

### Literature searching

Systematic search was performed through MEDLINE (OVID), Embase (OVID), Embase Classic, and the Cochrane Controlled Register of Trials (CENTRAL) and databases collecting publications from inception to April 2020. The following query terms were used: ‘electro-gastro*’ OR ‘electrogastro*’. There were no limits placed on date, language and patient age during the search. This non-restrictive search strategy was utilized to identify all potentially relevant studies using EGG.

### Inclusion and exclusion criteria

Studies were eligible for inclusion if they included adult patients aged 18 or over that were undergoing EGG. EGG was defined as any cutaneous recording of underlying gastric electrical activity. Studies which included EGG recordings for patients with FD, allowing for variable definitions of this disorder were included. Both subtypes of FD - epigastric pain syndrome and postprandial distress syndrome, as well as the legacy terms dysmotility-like and ulcer-like dyspepsia were analyzed. Studies which investigated treatments with appropriate control group or pre-treatment baseline recording were also included.

Methodological studies, case reports, technical notes, editorials, and commentaries were excluded. Studies which only had a healthy control, mixed or inadequately defined FD population were also excluded. All invasive recordings (i.e. serosal or mucosal) and high-resolution techniques were excluded as they represent distinct measurement tools. Studies in which there were surgical manipulation or alteration of gastrointestinal anatomy were excluded. Studies which investigated therapeutic interventions without an appropriate control group or baseline recordings were excluded. Non-English language manuscripts were excluded.

### Study selection and data extraction

All titles were screened for inclusion independently by two authors based on a pro forma. Two other independent authors further checked a random sample of 10% of these titles to ensure accurate capture. Discrepancies were discussed between authors with mediation by a third reviewer if required. A list of articles for full-text review was generated and data were extracted independently by four reviewers using a pro forma developed by the authors. Data extraction for each article was checked independently by at least two authors. In the case of discrepancies, these were discussed and resolved.

Information on study characteristics such as diagnostic criteria used, inclusion and exclusion criteria, and reference ranges, were extracted. Information on EGG methodology used was also extracted from each article, including: electrode number, electrode type, skin preparation, electrode placement method, and the EGG test protocol.

### Risk of bias assessment

The Quality Assessment of Diagnostic Accuracy Studies 2 (QUADAS-2) tool was used to assess study quality for all included studies ^13^. Domains for risk of bias assessed included: patient selection, index test, reference standard and participant flow and timing, with each domain graded as ‘low’, ‘high’, or ‘unclear’ risk of bias. Risk of bias assessments were checked independently by two authors, with remediation by a third reviewer in the event of a discrepancy.

### Outcomes

The clinical criteria used to diagnose FD in each article was recorded, along with any additional eligibility criteria used in each study. The primary outcomes were the percentage of recording duration where dominant power was in bradygastric (% bradygastria), normogastric (% normogastria) and tachygastric (% tachygastria) frequency ranges. Secondary outcomes included: dominant frequency (DF), dominant power (DP), DF instability coefficient (DFIC), DP instability coefficient (DPIC), power ratio and percentage coupling. The DFIC and DPIC were defined as the standard deviation (SD) of the DF (or DP) divided by the mean DF (or DP), over either the fasting or postprandial periods. Power ratio (PR) was defined as postprandial DP divided by the fasting DP. Slow waves in two channels were defined as coupled if the difference in their DF was <0.5 cycles per minute (cpm); the percentage of coupling between every pair of channels was then calculated and the average was taken. The prevalence of any EGG abnormality, as defined within studies, was also analyzed. The ratio of the meal response was determined by taking the postprandial value divided by the fasting value for each of the primary and secondary outcomes. The outcomes were recorded as either fasting, postprandial or meal-response ratio where applicable. If symptoms were evaluated throughout the test and considered alongside EGG metrics, any correlations including temporal correlations, were also analyzed.

### Statistical analysis

All statistical analyses were performed using R (version 4.0.2; R Foundation for Statistical Computing, Vienna, Austria). The *metacont* and *metaprop* packages were used ^14^.

Where studies reported continuous data as medians, mean estimates were calculated using the methods of Wan et al. ^15^ or Hozo et al. ^16^ as appropriate. Where EGG metrics were available from multiple channels, means and SD were combined as per the methods of Higgins et al ^17^. SD for meal-response ratios were calculated through error propagation ^18^. Data are presented as frequency (n) or percentage (%) for categorical outcomes and mean ± SD or median (range) for normal and non-normal continuous outcomes, respectively. In controlled studies, standardized mean differences (SMD) in EGG metric values were calculated and pooled via a random-effects model with the DerSimonian-Laird estimator for the between-study variance with 95% confidence interval (CI) ^19^. A random-effects model was chosen due to the expected between-study heterogeneity resulting from variability in experimental methods. Standardized mean differences (SMD) were used as the summary estimate in the meta-analyses due to expected variability in EGG metric definitions as per Holger et al. ^20^. SMD were back-transformed to the appropriate EGG-metric unit for % normogastria, bradygastria, tachygastria to aid interpretation ^21^. Heterogeneity was assessed using the I^2^ statistic. An I^2^ value of <25% was considered low heterogeneity, 25% to 75% was considered medium heterogeneity and >75% was considered high heterogeneity^22,23^. Publication bias and selective reporting of outcomes were assessed using Egger’s regression test and represented visually via funnel plots ^24^. A meta-analysis of EGG methodology and inconsistently reported outcomes was not conducted. These data were synthesised narratively or tabulated where appropriate. P-values <0.05 from the Egger’s regression test were considered statistically significant. In studies where the prevalence of abnormalities identified by EGG were reported, the pooled prevalence of EGG abnormalities were calculated using a random intercept logistic regression model (type of generalised linear mixed model), after log transformation ^25^. Clopper-Pearson 95% CIs were reported for individual studies. A 95%CI for the SMD which did not cross zero was considered statistically significant.

## Results

Database searching identified 3104 results of which 47 were included. Full search outcomes, including reasons for exclusion are shown in the PRISMA diagram (**Figure 1**). A detailed list of included studies can be found in **Table 1**.

**Figure 1.**
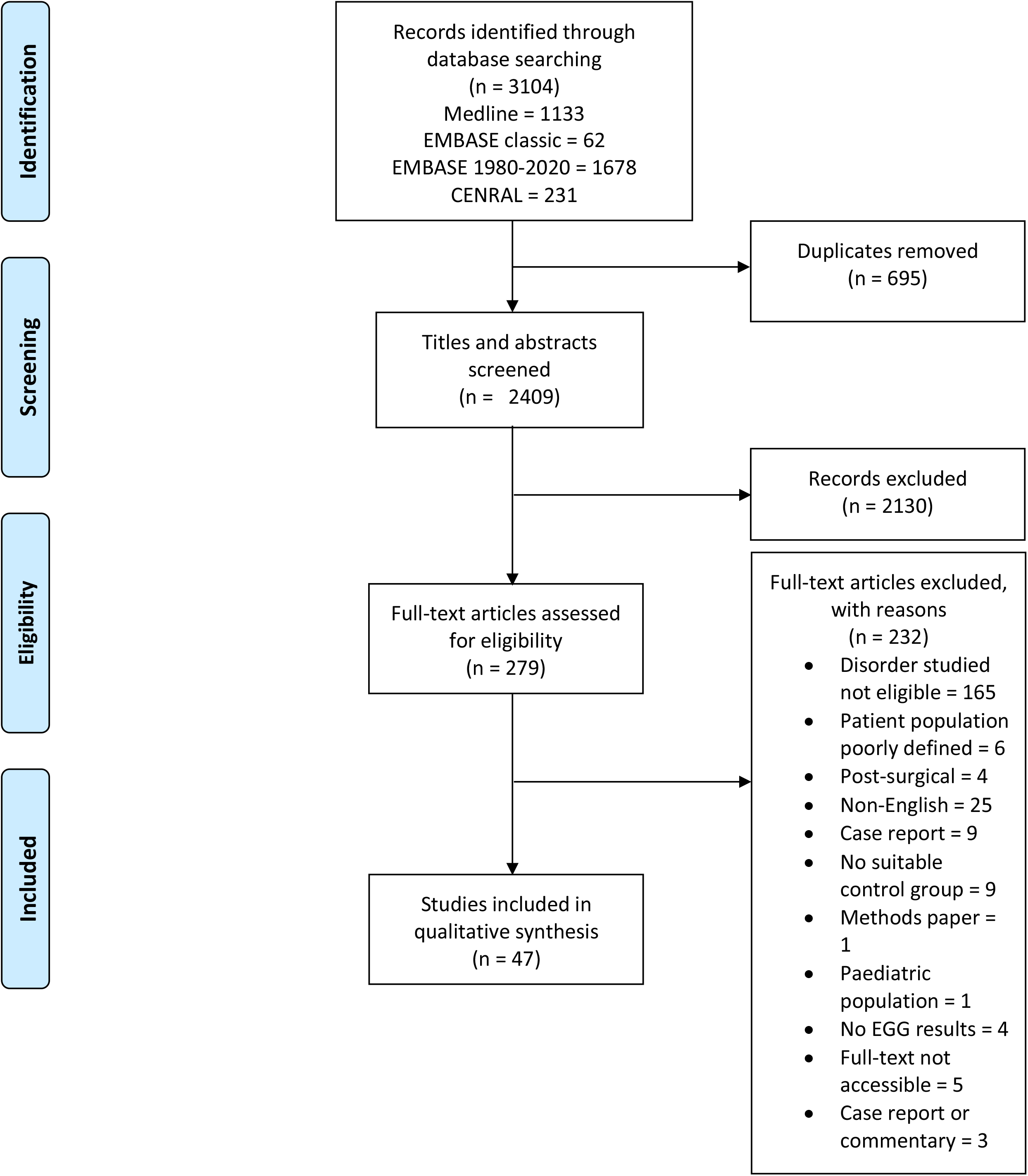
PRISMA flow diagram (EGG, electrogastrography)

In total, 1751 FD patients underwent recording of their gastric electrical activity using EGG, across the 47 included studies. Nearly twice as many females *vs* males (n = 1089 *vs* 499) were included, with sex not reported in 7 studies. FD not otherwise specified (NOS) was the most common patient population; reported in 35 studies (74.5%). The remaining 12 studies described the subtype of FD; 3 study cohorts classified dysmotility-like dyspepsia, 6 studies differentiated those with epigastric pain syndrome (EPS) and 8 studies differentiated those with postprandial distress syndrome (PDS). Further details on the included studies and patient characteristics are reported in **Table 1**.

All 47 studies were prospective in nature and most incorporated a control group (n = 36), with 4 of these articles being randomized controlled trials. The included articles spanned a 24-year period (1995 to 2019) and were conducted in a range of countries, most commonly in the United States (n = 14).

FD was diagnosed according to Rome III criteria in 14 studies, Rome II criteria in 12 studies, Rome I criteria in 6 studies and Talley et al.’s criteria in 3 studies ^26^. Eight studies did not specify the use of a published diagnostic criteria. Diagnostic and clinical criteria and study characteristics are reported in **Supplementary Table 1**.

### Clinical associations of gastric electrical activity identified by EGG

#### Fasting

Fasted FD patients spent 12% (SMD −0.74; 95%CI −1.22 - −0.25%, I^2^ = 76%) less time in normogastria compared to controls (**Figure 2**). FD patients also spent 5% (SMD 0.63; 95%CI 0.33 – 0.93%, I^2^ = 43%) more time in bradygastria and 4% (SMD 0.45; 95%CI 0.12 – 0.76%, I^2^ = 38%) more time in tachygastria compared to controls (**Figure 2**). The fasted DF was not significantly different between the FD group and controls (SMD −0.31; 95%CI −0.69 - 0.07, I^2^ = 71%) (**Appendix 1**). Similarly, the fasted DP was also not significantly different between the FD group compared to controls (SMD 0.34; 95%CI −0.50 - 1.17, I^2^ = 80%) (**Appendix 1**). Nor was DFIC (SMD −0.37; 95%CI −4.54 - 3.79, I^2^ = 96%) (**Appendix 1**).

**Figure 2.**
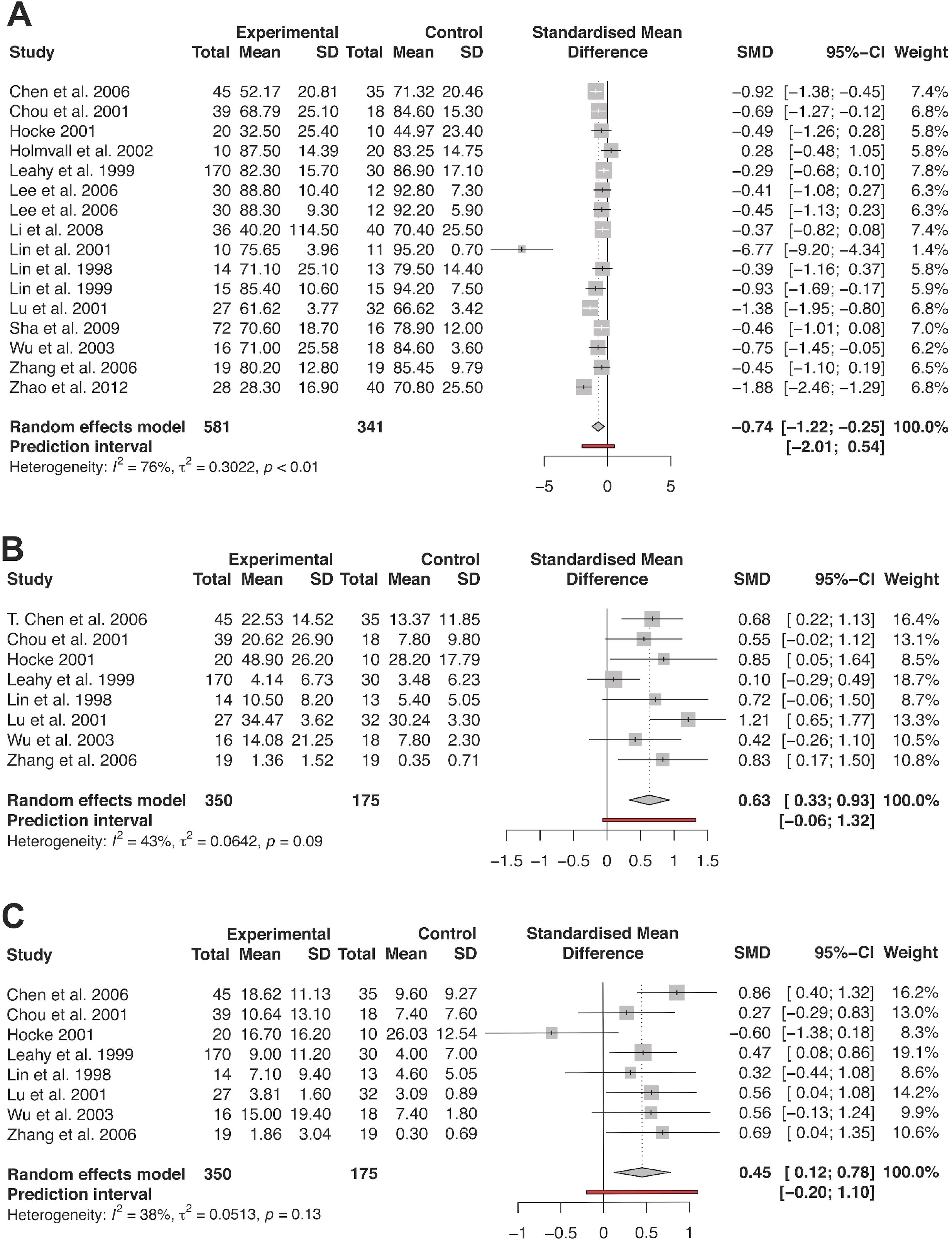
Forest plots of fasting percentage frequency electrogastrography metrics; A: fasting percentage normogastria, B: fasting percentage bradygastria, C: fasting percentage tachygastria.

#### Postprandial

Postprandial FD patients spent 13% less time in normogastria (SMD 0.86; 95%CI −1.35 - - 0.37%, I^2^ = 85%) compared to controls (**Figure 3**). The increased time spent in bradygastria (SMD 0.55; 95%CI −0.03 – 1.14 %, I^2^ = 81%), and tachygastria (SMD 0.38; 95%CI −0.45 – 1.21%, I^2^ = 88%) compared to controls postprandially did not reach significance (**Figure 3**). The DF was not significantly different between the FD group and controls (SMD −0.13; 95% CI −0.51 - 0.24, I^2^ = 75%) (**Appendix 1**). Similarly, the DP was also not significantly different between the FD group compared to controls (SMD −0.90; 95% CI −3.43 - 1.63, I^2^ = 94%), nor was the DFIC (SMD 0.73; 95% CI −0.04 - 1.50, I^2^ = 88%) (**Appendix 1**).

**Figure 3.**
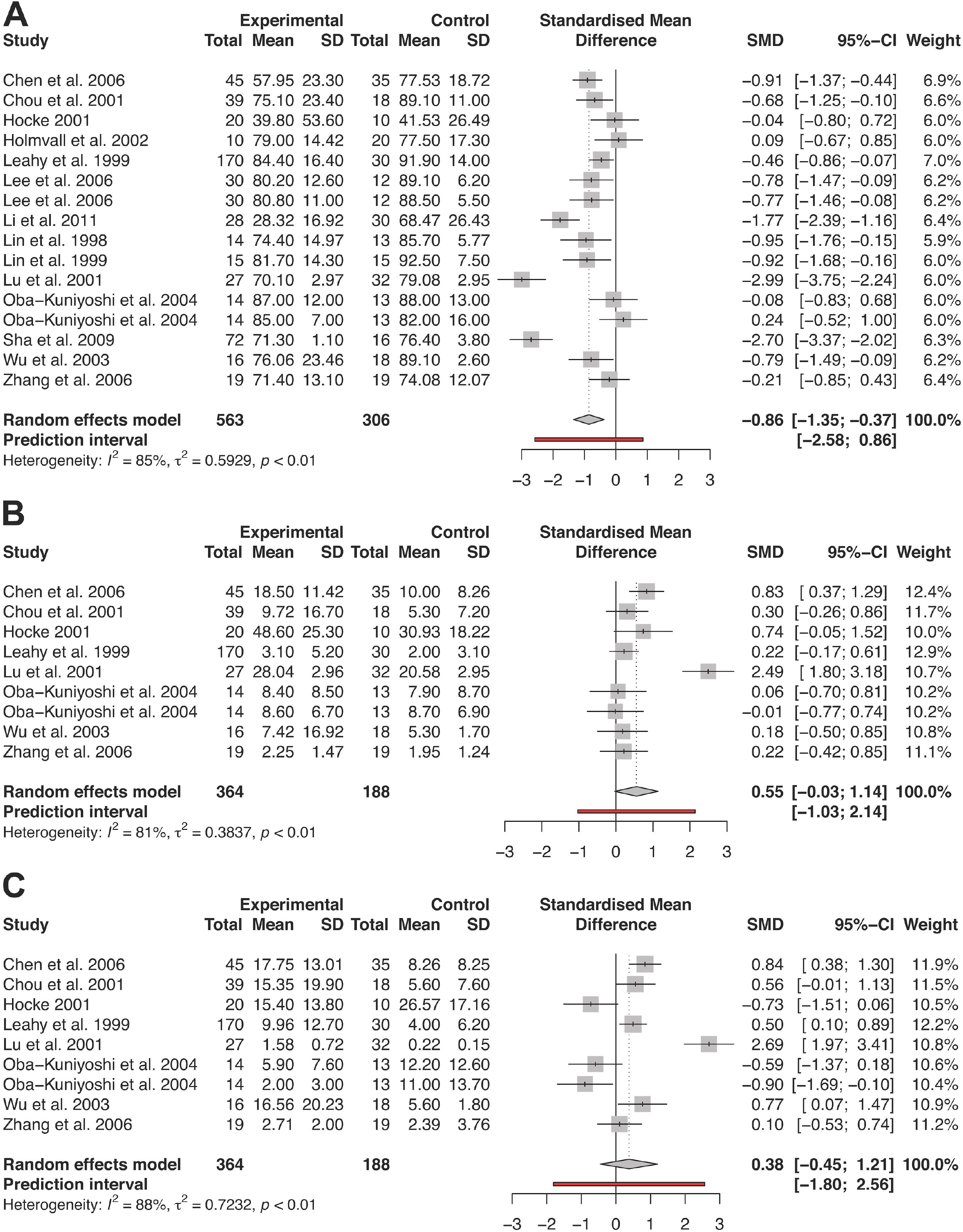
Forest plots of postprandial percentage frequency electrogastrography metrics; A: postprandial percentage normogastria, B: postprandial percentage bradygastria, C: postprandial percentage tachygastria.

#### Meal Response

The PR was not significantly different between FD patients and controls (SMD −0.17; 95% CI −0.83 - 0.48, I^2^ = 88%). These results were not significantly different in a subgroup analysis of studies excluding manually calculated PR (i.e. postprandial divided by fasting DP) (**Appendix 1**). One study reported postprandial power change (i.e. the difference between postprandial and fasting DP), which was not significantly different between FD patients and controls ^8^.

The meal response ratio of % normogastria was not significantly different between FD patients and controls (SMD 0.07 95% CI −0.14 - 0.28, I^2^ = 34%) nor the ratio of % bradygastria (SMD −0.04 95% CI −0.23 - 0.14, I^2^ = 0%), or % tachygastria (SMD 0.30 95% CI −0.38 - 0.98, I^2^ = 82%). The meal response ratio of DF was also not significantly different between FD patients and controls (SMD 0.06 95% CI −0.08 - 0.21, I^2^ = 0%), nor the meal response ratio of DFIC (SMD 0.41 95% CI −0.35 - 1.17, I^2^ = 84%).

#### Coupling

FD patients trended towards lower percentage slow wave coupling while fasted (SMD −1.47 95% CI −3.72 – 0.79, I^2^ = 96%). Postprandial percentage slow wave coupling, and meal response percentage coupling ratio were not meta-analysed due to low number of studies (≤2). Coupling metrics can be found in **Appendix 1**. Zhang et al. utilized a different definition for coupling, where the difference in DF between slow waves had to be <0.2 cpm and only slow waves within normogastric DFs were included ^26^. This study also found negative correlations between percentage slow wave coupling and 1- and 2-hour retention rate on gastric emptying tests.

Forest and funnel plots for all primary and secondary outcomes can be found in the **Supplementary Appendix 1**.

#### Pooled Prevalence of EGG abnormalities

The proportion of FD patients with any EGG abnormality was reported in 24 studies. The pooled prevalence of any EGG abnormality was 51% (95% CI 45%-58%, I^2^ = 75%), **Supplementary Figure 1**.

#### Symptom correlations

Nineteen studies commented on the correlation between FD patient symptoms and EGG metrics. Of these, the majority (63%) reported no correlations between EGG and FD symptoms. Two studies suggested abnormal EGG was associated with higher nausea rates ^27^, and another suggested that those with abnormal EGG had a higher Glasgow dyspepsia scale ^28^. One study found a correlation between upper abdominal discomfort and anorexia with abnormal EGG, but not nausea, vomiting, early satiety and abdominal discomfort. Two studies showed a correlation between dyspepsia symptoms in general and abnormal EGG ^29,30^. Seven studies investigated temporal correlations between EGG metrics and symptoms, of which three reported no correlation ^31,32^. The other four studies found EGG abnormalities correlated with abnormal gastric emptying ^26,33–35^.

Among the few studies that separated EGG results based on FD subtypes of EPS or PDS, Rudnicki et al. found no differences in the DF, % bradygastria or tachygastria or % arrhythmia between the two subtypes (all p>0.05) ^37^. Russo et al. found similar results, however noted PDS patients had higher DFIC and a lower DP compared to EPS (p=0.03 for both) ^38^.

### EGG methodology

From 44 (93.6%) studies, a mode of three electrodes (range 2-6) were used to record surface electrical activity. Silver-silver chloride electrodes are used as standard in EGG and 12 (26%) studies described the use of electroconductive gel. Of 42 (89%) studies which reported the type of recorded signal taken, 18 (43%) took bipolar recordings while the remaining 24 (57%) were unipolar. Of 38 studies where skin preparation was described, some form of abrasion, exfoliation or cleaning occurred prior to application of electrodes in 36 (95%) studies, and 18 (47%) studies reported shaving hair to improve conductance. Electrode placement was very heterogeneous, but most studies (40/43, 93%) standardised electrode placement based on defined anatomical landmarks. Only two studies (4%) used ultrasound guided electrode placement and five studies either did not specify or used vague placement criteria. Nineteen studies (40%) had patients fast overnight before EGG recordings, 2 studies had patients fast for 12 hours, 7 for 8 hours, 6 for 6 hours and 3 for 2 hours. Pre-recording fasting duration was not specified in 10 studies. Most studies (n = 38) used a meal stimulus, while 5 studies employed a water load test, and 2 studies only included passive EGG recordings. Meals and caloric intake varied significantly. Duration of EGG recordings varied from 30 minutes to 24 hours, commonly with 30 minutes (21, 46%) of pre-meal recordings followed by 30 (14, 33%) or 60 (14, 33%) minutes of post-prandial recordings. Further EGG details can be found in **Table 3**. Most studies also stopped medications prior to the EGG study, however this was not stated in 10 studies, and in three studies medications were not stopped ^39–41^.

Of the 43 studies which stated the range of slow wave frequencies defined as normogastria, 34 (79%) used 2 cpm as the lower limit, and 36 (84%) used 4 cpm as the upper limit. A minority of studies used a lower limit of 2.4^42,43^, 2.5^28,31,44–48^ or 2.6^32^ and an upper limit of 3.5^47,48^, 3.7^32,42,43^, or 3.75^28,31,44^.

### Risk of bias

When assessing patient selection methods, 25 (53%) studies had an unclear risk of bias, 12 (26%) studies had a high risk of bias and 10 (21%) studies had a low risk of bias. With regards to the index test (EGG), there were 16 (34%) studies with unclear risk of bias, 17 (36%) studies with high risk of bias and 14 (30%) studies with low risk of bias When assessing the use of the reference standard (i.e. the gold standard diagnostic tool) in included studies, 5 (11%) studies had an unclear risk of bias, only 1 (2%) was associated with a high risk of bias, and the majority, 41 (87%), had a low risk of bias. Similarly, in the domain of patient flow and timing, 7 (15%) studies had an unclear risk of bias, 3 (6%) had a high risk of bias and 37 (79%) studies had a low risk of bias. Further QUADAS-2 assessment details are reported in **Supplementary Table 3** and **Figure 4**.

**Figure 4.**
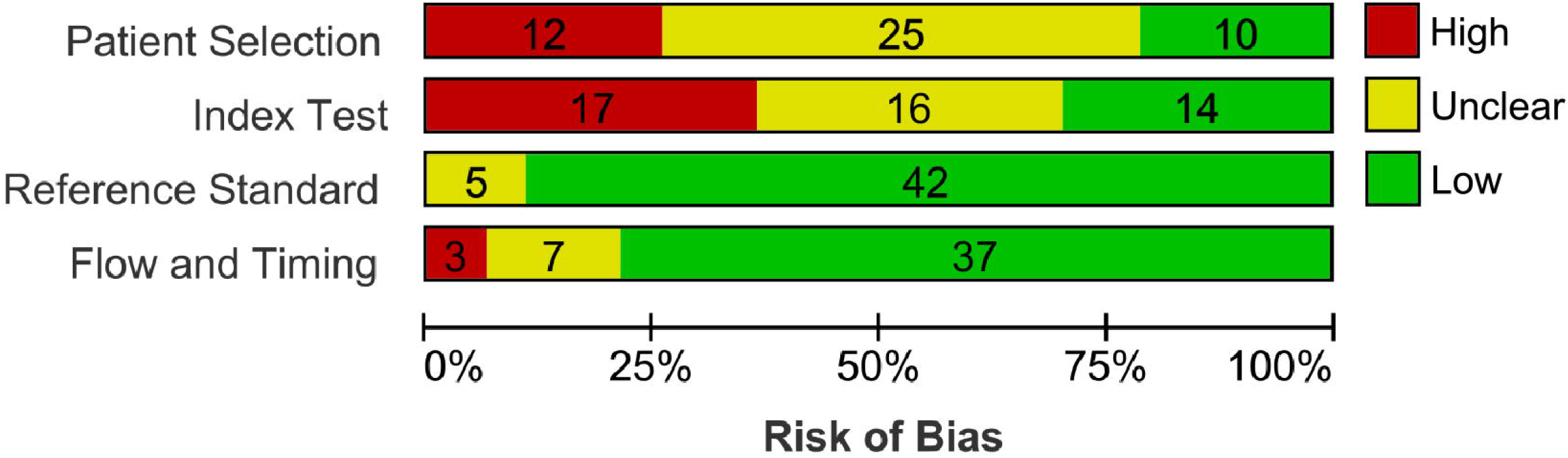
QUADAS-2 risk of bias assessment

There was a relatively low degree of publication bias on inspection of funnel plots. All Egger’s test p values were >0.05 except for the postprandial DF and fasting DFIC. Further details can be found in **Appendix 1**.

## Discussion

This is a comprehensive systematic review and meta-analysis of EGG abnormalities in FD, compiling and synthesizing a substantial existing literature as a foundation for future studies. Several frequency-based EGG-metrics were revealed to show strong and consistent associations in FD. In the fasted state, FD patients spent 12% less time in normogastria, 5% more time in bradygastria, and 4% more time in tachygastria compared to controls. After a meal, FD patients spent 13% less time in normogastria. Power metrics and meal response ratios were not significantly different between FD patients and controls, and there were few and inconsistent correlations between EGG metrics and symptoms.

A key finding of this review was that FD patients spend substantially shorter periods in normal gastric rhythms. It was noteworthy that these frequency abnormalities were consistent across a large number of studies, as well as across fasting and fed states, indicating that they may signal an important and relatively under-appreciated feature of FD pathophysiology. The mechanism of these frequency abnormalities is poorly understood, and warrants further research. Multiple factors are known to have chronotropic influence on gastric slow waves, including intrinsic interstitial cells of Cajal (ICC) pacemaker activity, ion channel behaviours, sensitivity to stretch, as well as autonomic, enteric nervous system, hormonal, and paracrine factors ^49^, but which of these are responsible for the features summarized here in FD patients is currently unresolved. While damage and loss of ICC has been extensively reported in gastroparesis and chronic nausea and vomiting syndromes, and may explain disrupted slow wave initiation and conduction in those disorders ^9,10,50^, such findings are not described in FD. The increasing awareness of brain-gut interactions, mucosal inflammation, and increased sensitivity to gastric distention in FD all present possible paths of enquiry to link with electrophysiological abnormalities ^2,51,52^.

Tack et al. suggested five criteria for the plausibility of putative pathophysiological mechanisms in functional gastrointestinal disorders ^53^, and it is of interest to apply these criteria to this review. Evidence is found in support of Criteria 1, in that a substantial subset of FD patients (up to 51%) have EGG abnormalities. With regards to Criteria 2, temporal correlations between EGG abnormalities and FD symptoms were inconsistent at best in the limited available data. While the present EGG literature also sparsely investigated a correlation between the severity of EGG abnormalities and severity of FD symptoms (Criteria 3), these associations have recently been suggested in a high-resolution study ^8^. Correlations between EGG metrics and specific FD symptoms such as postprandial fullness, epigastric pain and burning have not been well defined, although a small number of studies have shown associations with early satiety, bloating and abdominal pain ^8,54^. Future studies could continue to address these associations and the remaining plausibility criteria to further resolve the mechanistic relationship between aberrant gastric electrophysiology and FD.

It is highly likely that the dysrhythmias identified in FD cause disordered gastric motility ^55–57^. Normal gastric slow wave coupling (entrainment) fundamentally depends upon the presence of functioning frequency gradients, and frequency disturbances are therefore associated with aberrant patterns of propagation including rapid circumferential conduction and retrograde activation ^49,58^. Gastric motility is also known to be suppressed during dysrhythmias, particularly tachygastria ^59,60^, providing one plausible link between electrophysiological abnormalities and the antral hypomotility and delayed gastric emptying that has been described in subsets of FD patients ^61,62^. However, it is also now understood that frequency alone is a poor proxy for the normality of slow wave patterns, because propagation abnormalities also commonly occur at normal frequencies in humans ^63^. No studies in this review employed more than 6 electrodes for EGG recordings, meaning they were unable to reliably measure spatial metrics such as propagation patterns and spatial discoordination which may also be relevant to gastric dysmotility.

As well as a lack of spatial resolution, other limitations of EGG highlighted by this review include inconsistencies in electrode placement, data acquisition systems, recording methods, metrics, and study protocols. In particular, variations in meals used and duration of recordings are likely to affect results. In several instances, there was a high degree of statistical heterogeneity, likely accounted for by variabilities in methodologies and patient cohorts. However, the broadly similar normative frequency ranges and use of a random-effects meta-analysis mitigated this issue for the primary outcomes. Pooled meta-analyses generally also showed low degrees of publication bias on visual inspection of funnel plots and Egger’s regression test, with the exception of fasting DFIC and postprandial DF. However, over three-quarters of the included studies had either high or unclear risk of bias with regards to patient selection, study design, and appropriateness of patient exclusions. Future studies should ideally blind the interpretation of EGG results and employ pre-specified reference ranges. In addition, traditional EGG techniques as used in this review are known to have a low signal-to-noise ratio such that they are prone to contamination by artefacts ^64,65^. These artefacts could have affected the pooled data used in the review, but it is notable that electrophysiological abnormalities were nevertheless robustly identified in this meta-analysis.

Many of the limitations observed above have also severely dampened clinical enthusiasm for using EGG in the diagnosis of gastroduodenal disorders including FD ^66^, such that it is now rarely used outside of interested specialist centres. New techniques that are standardized, user-friendly and that offer substantially more reliable actionable biomarkers would be necessary to restore clinical enthusiasm ^11^. The emergence of next-generation high-resolution approaches to gastric electrophysiology, and notably ‘body surface gastric mapping’ ^63^ are indicating potential, including due to the introduction of novel non-invasive biomarkers such as retrograde propagation that may be most relevant to FD symptoms ^8,67^. The use of these techniques is anticipated to grow in the coming years, and this systematic review, which is based on a vast volume of research experience using traditional EGG, will provide an essential foundation to inform their application to FD.

In conclusion this systematic review and meta-analysis found FD patients spent significantly less time in normogastria in the fasted and fed states compared to controls. The aberrant electrophysiology identified in FD by EGG is shown to be consistent across a large number of studies; it may be under-appreciated, and remains unexplained. Given the high prevalence and impact of FD, and the lack of effective therapies, these abnormalities warrant further investigation by more advanced methods, including into their responsible mechanisms, consequences for motility, relevance to symptom genesis, and therapeutic significance.

## Data Availability

This report contains no primary data.

## Figure Legends (order they appear in manuscript)

**Table 1**. Characteristics of included studies investigating functional dyspepsia

**Supplementary Table 1**. Clinical characteristics of functional dyspepsia in included studies

**Supplementary Figure 1.**
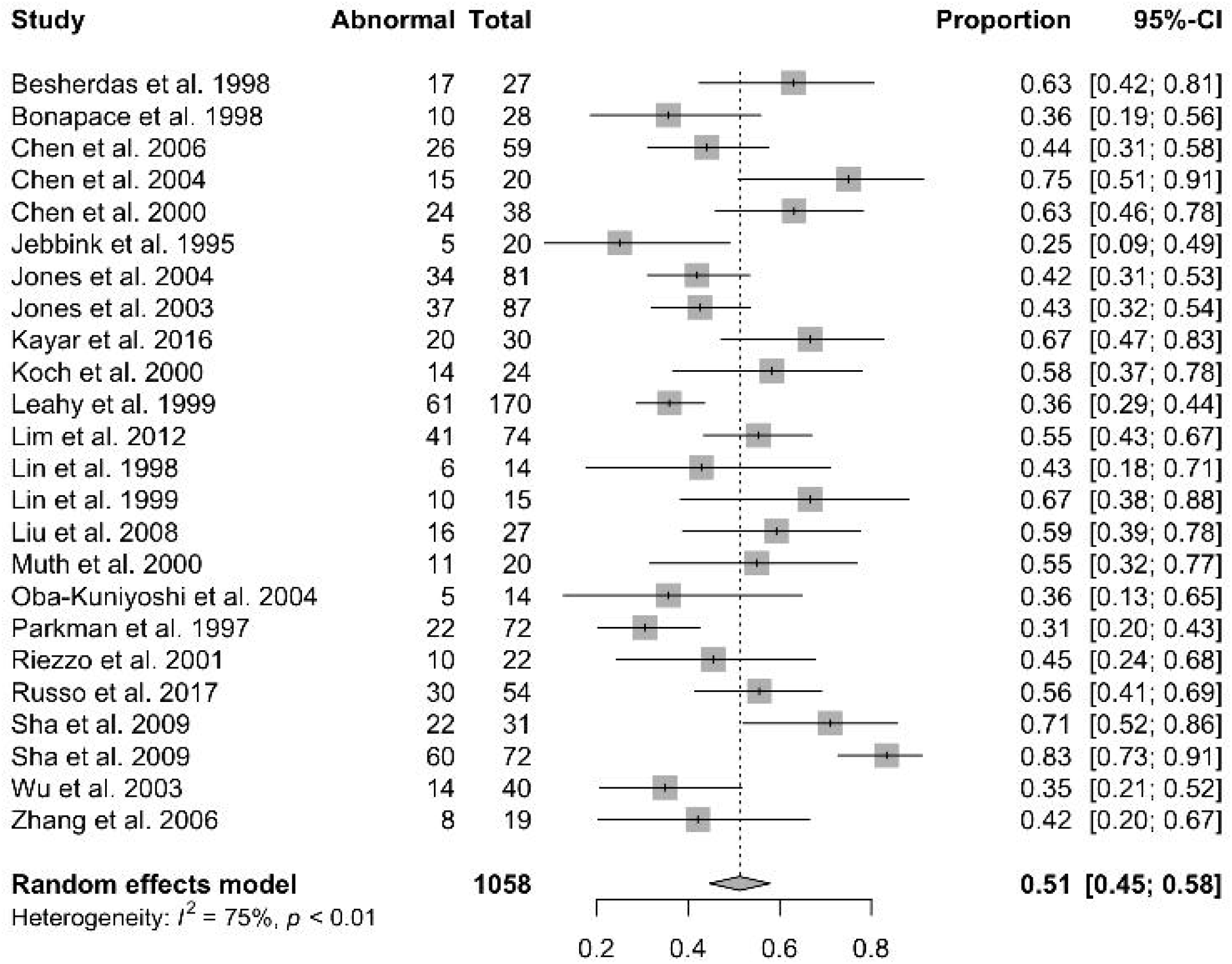
Forest plot of electrogastrography abnormalities in functional dyspepsia patients

**Supplementary Table 2**. Electrogastrography parameters in functional dyspepsia patients

**Supplementary Table 3**. QUADAS-2 risk of bias assessment of all included studies

