## Appendix for "Clinical Associations of Functional Dyspepsia with Gastric Dysrhythmia on Electrogastrography: A Comprehensive Systematic Review and Meta-Analysis"

| **Reference** | **Year** | **Geographic Location** | **Disorder** | **Number of participants in disorder group above** | **Male and female participants, express as "M, F"** | **Study Design** |
| --- | --- | --- | --- | --- | --- | --- |
| Al Kafee et al. | 2018 | Turkey | Functional dyspepsia (NOS) | 30 | 6M, 24F | Prospective, Non-randomised, controlled study |
| Besherdas et al. | 1998 | United Kingdom | Functional dyspepsia (NOS) | 27 | Not reported | Prospective, Non-randomised, non-controlled study |
| Bonapace et al. | 1998 | United States | Functional dyspepsia (NOS) | 28 | 6M, 22F | Prospective, Non-randomised, controlled study |
| Chacon et al. | 2009 | Chile | Functional dyspepsia (NOS) | 42 | 6M, 36F | Prospective, Non-randomised, controlled study |
| C. Chen et al. | 2006 | Taiwan | Functional dyspepsia (NOS) | 59 | 13M, 46F | Prospective, Non-randomised, controlled study |
| Chen et al. | 2004 | Taiwan | Functional dyspepsia (NOS), GORD | 20 | 8M, 12F | Prospective, Non-randomised, non-controlled study |
| Chen et al. | 2000 | United States | Functional dyspepsia (NOS) | 38 | 15M, 23F | Prospective, Randomised, controlled study |
| T. Chen et al. | 2006 | Taiwan | Functional dyspepsia (NOS) | 45 | 21M, 24F | Prospective, Non-randomised, controlled study |
| Chou et al. | 2001 | Taiwan | Functional dyspepsia (NOS) | 39 | 15M, 24F | Prospective, Non-randomised, controlled study |
| De Giorgi et al. | 2013 | Italy | Functional dyspepsia (PDS) | 25 | 11M, 14F | Prospective, Non-randomised, controlled study |
| Gharibans et al. | 2019 | United States | Functional dyspepsia (PDS, Functional dyspepsia (EPS), Functional dyspepsia (PDS and EPS) | 7 | 3M, 4F | Prospective, Non-randomised, controlled study |
| Hocke et al. | 2001 | Switzerland | Functional dyspepsia (NOS) | 20 | 6M, 14F | Prospective, Non-randomised, controlled study |
| Holmvall et al. | 2002 | Sweden | Functional dyspepsia (NOS) | 10 | 1M, 9F | Prospective, Non-randomised, controlled study |
| Jebbink et al. | 1995 | Netherlands | Functional dyspepsia (NOS) | 20 | 8M, 12F | Prospective, Non-randomised, controlled study |
| Ji et al. | 2014 | United States | Functional dyspepsia (PDS) | 28 | 15M, 13F | Prospective, Randomised, controlled study |
| Jones et al. | 2004 | United States | Functional dyspepsia (NOS) | 81 | 20M, 61F | Prospective, Non-randomised, controlled study |
| Jones et al. | 2003 | United States | Functional dyspepsia (NOS) | 87 | 21M, 66F | Prospective, Non-randomised, controlled study |
| Jung et al. | 2012 | South Korea | Functional dyspepsia (PDS), Functional dyspepsia (EPS) | 13 | 4M, 9F | Prospective, Non-randomised, non-controlled study |
| Kayar et al. | 2016 | Turkey | Functional dyspepsia (NOS) | 30 | 6M, 24F | Prospective, Non-randomised, controlled study |
| Kim et al. | 2012 | South Korea | Functional dyspepsia (PDS), Functional dyspepsia (EPS) | 22 | 0M, 22F | Prospective, Non-randomised, controlled study |
| Koch et al. | 2000 | United States | Functional dyspepsia (dysmotility-like) | 24 | 7M, 17F | Prospective, Non-randomised, controlled study |
| Leahy et al. | 1999 | United Kingdom | Functional dyspepsia (NOS) | 170 | 65M, 105F | Prospective, Non-randomised, controlled study |
| Lee et al. | 2006 | South Korea | Functional dyspepsia (NOS) | 30 | 13M, 17F | Prospective, Non-randomised, controlled study |
| Li et al. | 2011 | China | Functional dyspepsia (NOS) | 28 | Not reported | Prospective, Non-randomised, controlled study |
| Li et al. | 2008 | China | Functional dyspepsia (NOS) | 36 | Not reported | Prospective, Non-randomised, controlled study |
| Lim et al. | 2012 | Korea | Functional dyspepsia (NOS) | 74 | 26M, 48F | Prospective, Non-randomised, non-controlled study |
| Lin et al. | 2001 | United States | Functional dyspepsia (NOS) | 10 | 2M, 8F | Prospective, Non-randomised, controlled study |
| Lin et al. | 1998 | United States | Functional dyspepsia (NOS) | 14 | 4M, 10F | Prospective, Non-randomised, controlled study |
| Lin et al. | 1999 | United States | Functional dyspepsia (NOS) | 15 | 2M, 13F | Prospective, Non-randomised, controlled study |
| Liu et al. | 2008 | China | Functional dyspepsia (NOS) | 27 | 9M, 18F | Prospective, Randomised, controlled (sham) study |
| Lu et al. | 2001 | Taiwan | Functional dyspepsia (NOS) | 27 | 10M, 17F | Prospective, Non-randomised, controlled study |
| Muth et al. | 2000 | United States | Functional dyspepsia (dysmotility-like) | 20 | 5M, 15F | Prospective, Non-randomised, non-controlled study |
| Oba-Kuniyoshi et al. | 2004 | Brazil | Functional dyspepsia (dysmotility-like) | 14 | 5M, 9F | Prospective, Non-randomised, controlled study |
| Park et al. | 2013 | Korea | Functional dyspepsia (PDS, Functional dyspepsia (EPS), Functional dyspepsia (PDS and EPS) | 100 | 47M, 53F | Prospective, Randomised, controlled study |
| Parkman et al. | 1997 | United States | Functional dyspepsia (NOS) | 72 | 12M, 60F | Prospective, Non-randomised, non-controlled study |
| Pfaffenbach et al. | 1997 | Germany | Functional dyspepsia (NOS) | 25 | 8M, 17F | Prospective, Non-randomised, controlled study |
| Riezzo et al. | 2001 | Italy | Functional dyspepsia (NOS) | 22 | 7M, 15F | Prospective, Non-randomised, controlled study |
| Rudnicki et al. | 2009 | Poland | Functional dyspepsia (PDS) | 6 | Not reported | Prospective, Non-randomised, non-controlled study |
| Rudnicki et al. | 2009 | Poland | Functional dyspepsia (EPS) | 22 | Not reported | Prospective, Non-randomised, non-controlled study |
| Russo et al. | 2017 | Italy | Functional dyspepsia (PDS), Functional dyspepsia (EPS) | 54 | 4M, 50F | Prospective, Non-randomised, non-controlled study |
| Sha et al. | 2009 | United States | Functional dyspepsia (NOS) | 31 | 8M, 23F | Prospective, Non-randomised, non-controlled study |
| Sha et al. 2 | 2009 | United States | Functional dyspepsia (NOS) | 72 | 23M, 49F | Prospective, Non-randomised, controlled study |
| Shiotani et al. | 2002 | Japan | Functional dyspepsia (NOS) | 64 | 26M, 38F | Prospective, Non-randomised, non-controlled study |
| Van Der Voort et al. | 2003 | Germany | Functional dyspepsia (NOS) | 16 | Not reported | Prospective, Non-randomised, controlled study |
| Wu et al. | 2003 | Taiwan | Functional dyspepsia (NOS) | 40 | 16M, 24F | Prospective, Non-randomised, controlled study |
| Qu et al. | 2010 | China | Functional dyspepsia (NOS) | 20 | 8M, 12F | Prospective, Non-randomised, non-controlled study |
| Zhang et al. | 2006 | China | Functional dyspepsia (NOS) | 19 | 7M, 12F | Prospective, Non-randomised, controlled study |
| Zhao et al. | 2012 | China | Functional dyspepsia (NOS) | 28 | Not reported | Prospective, Non-randomised, controlled study |
